# Dying From COVID-19 or With COVID-19: A Definitive Answer Through a Retrospective Analysis of Mortality in Italy

**DOI:** 10.1101/2021.12.22.21268212

**Authors:** Alessandro Rovetta, Akshaya Srikanth Bhagavathula

## Abstract

**Background:** COVID-19 mortality was associated with several reasons, including conspiracy theories and infodemic phenomena. However, little is known about the potential endogenous reasons for the increase in COVID-19 associated mortality in Italy.

**Objective:** This study aimed to search the potential endogenous reasons for the increase in COVID-19 mortality recorded in Italy during the year 2020 and evaluate the statistical significance of the latter.

**Methods:** We analyzed all the trends in the timelapse 2011-2019 related to deaths by age, sex, region, and cause of death in Italy and compared them with those of 2020. Ordinary least squares (OLS) linear regressions and ARIMA (p, d, q) models were applied to investigate the predictions of death in 2020 as compared to death reported in the same year. Grubbs and Iglewicz-Hoaglin tests were used to identify the statistical differences between the predicted and observed deaths. The relationship between mortality and predictive variables was assessed using OLS multiple regression models.

**Results:** Both ARIMA and OLS linear regression models predicted the number of deaths in Italy during 2020 to be between 640,000 and 660,000 (95% confidence intervals range: 620,000 – 695,000) and these values were far from the observed deaths reported (above 750,000). Significant difference in deaths at national level (P = 0.003), and higher male mortality than women (+18% versus +14%, P < 0.001 versus P = 0.01) was observed. Finally, higher mortality was strongly and positively correlated with latitude (R = 0.82, P < 0.001).

**Conclusions:** Our findings support the absence of historical endogenous reasons capable of justifying the increase in deaths and mortality observed in Italy in 2020. Together with the current knowledge on the novel coronavirus 2019, these findings provide decisive evidence on the devastating impact of COVID-19 in Italy. We suggest that this research be leveraged by government, health, and information authorities to furnish proof against conspiracy hypotheses. Moreover, given the marked concordance between the predictions of the ARIMA and OLS regression models, we suggest that these models be exploited to predict mortality trends.

## Introduction

### Background

SARS-CoV-2 is a new beta coronavirus first identified in December 2019 in Wuhan, China. The related pathology, called COVID-19, has raged worldwide, claiming millions of victims and throwing economic and health systems into severe crises. In such a dramatic scenario, Europe is one of the most affected areas: as of December 2021, it accounts for over 30% of global official deaths (i.e., approximately 1,600,000) [1]. Because the risk factors are multiple, including environmental conditions, pollution, age, gender, ethnicity, crowding, poverty, and medical comorbidities, mortality varies substantially from country to country as well as intra-nationally [2–4]. Indeed, the daily deaths’ peaks per million inhabitants ranged from 1 (Ukraine) to over 40 (Belgium), with a median of 3.5 (IQR: 2 – 13) [4]. The first European nation to suffer the devastating effects of COVID-19 was Italy, with mortality peaks much higher than the European median (over 15). In particular, the regions of Northern Italy - especially the provinces of Bergamo and Brescia - faced a harsh first wave, reaching the highest number of deaths globally [1, 5]. To date, despite a substantial reduction in mortality thanks to a massive vaccination campaign, Italy is still the second European country for COVID-19 official deaths [1, 6]. Nonetheless, the debate over COVID-19 mortality has been intense during the pandemic. In the early stages, given the low testing capabilities, the calculation of mortality was subject to numerous uncertainties, which led to both overestimates and underestimates. For this reason, the researchers focused their efforts on comparing the 2020 data with the historical death series [7].

Ordinary least squares (OLS) regressions are among the most adopted model by scientists due to their simplicity and efficacy. Specifically, OLS multiple and simple regressions have often been used to predict the course of COVID-19 cases and deaths, both individually and in conjunction with other epidemiological models such as Susceptible-Infected-Recovered (SIR) [8–10]. This literature showed that linear regressions are valuable short-term forecasting tools when the necessary assumptions are satisfied. However, it is not unusual for requests such as normality of the residuals or homoskedasticity to be violated when dealing with actual epidemiological data. In these cases, the use of corrective procedures or alternative models should be considered. Among the latter, ARIMA (autoregressive integrated moving average) and SARIMA (ARIMA + seasonal component) models have shown excellent predictive capabilities. In particular, a recent study by Abolmaali and Shirzaei demonstrated that the ARIMA approach could outperform other classical models such as logistic function, linear regression, and SIR [11]. Similar findings were obtained by Alabdulrazzaq et al., who proved that the accuracy of the prediction of COVID-19 spread provided by their ARIMA model was both appropriate and satisfactory [12].

Despite above 135,000 official deaths nationwide, some Italian conspiracy movements argue that COVID-19 is a non-dangerous disease and that these numbers have been deliberately exaggerated [13]. Unfortunately, it was not uncommon even for eminent Italian scientists or other prominent personalities to have recklessly downplayed the risks of COVID-19 or favored the spread of fake news [14, 15]. Thus, the infodemic question “Dead from COVID or with COVID?” soon filled social networks [13]. Indeed, such a question arises from the hypothesis that COVID-19 was non-causally correlated with the deaths recorded in Italy in 2020.

Based on the above premise, this study aimed to estimate the difference between the observed and predicted number of deaths in Italy during 2020. In particular, we modelled all mortality trends by cause of death, sex, and age group from 2011 to 2019, predicting the best values for 2020. By doing so, causal evidence will be provided on the impact of a non-endogenous mortality factor, such as COVID-19. The results of this paper have epidemiological and infodemiological relevance since i) two models widely adopted by the scientific community such as OLS linear regression and ARIMA are compared, ii) to the best of our knowledge, this is the most detailed historical and forecasting survey regarding mortality in Italy, iii) an estimate of the statistical significance of the increase in mortality in Italy during 2020 is provided.

## Methods

### Data collection

For this study, we used data from the national agencies and portals of demographic and statistical research, Italy (details and references are provided below). Specifically, the annual number of deaths (including deaths by sex and age groups), deaths per causes of deaths (including deaths by sex groups), and mortality (including mortality by sex and age groups) were extracted from the platforms and annual reports of the National Institute of Statistics (*ISTAT*) and National Health Observatory for the years 2011-2020 [16–18]. Demographic data (i.e., population number per age group, population number and density, and per region) were gathered from *Tuttitalia.it*. [19, 20]. This portal contains all ISTAT demographic information relating to municipalities, provinces, and regions. While the investigated period ranged from 2011 to 2020, causes of death statistics were available until 2017 as the official evaluation process takes three years [17]. More details on the data collection process are described in Supplementary file 1, section 1.

### Procedure and statistical analysis

We modeled regional trends in annual deaths from 2011 to 2019 through ordinary least squares (OLS) linear regression. The standard assumptions for this regression model were automatically checked by the *XLSTAT Linear Regression* tool (default settings) [21]. However, a qualitative graphic check was also performed to observe any non-normality of the residuals, heteroskedasticity, or outliers. Goodness-of-fit measures have been reported in the results section and supplementary tables. Calling *Y* the observed values and *Y’* the values predicted by the interpolating line, we normalized the two variables by the following operation: *y’=Y’/max(Y;Y’)* and *y=Y/max(Y;Y’)*. Then, we calculated the residuals as δ*=y’-y*. Calling Δ*\** the residual dataset from 2011 to 2019 and Δ the residual dataset from 2011 to 2020, we performed the following steps: i) we evaluated the distribution normality of Δ*\** through the Shapiro-Wilk test plus Q-Q and Box plots, ii) we evaluated the distribution normality of Δ through the Shapiro-Wilk test plus Q-Q and Box plots, iii) through Grubbs and Iglewicz-Hoaglin tests plus a graphical check, we searched for outliers in Δ*\**, and iv) through Grubbs and Iglewicz-Hoaglin tests plus a graphical check, we searched for outliers in Δ. In this way, we assessed whether the residual δ*_2020* belonged to the Δ*\** distribution (ie, if there was an anomaly in national and regional deaths during 2020 compared to 2011-2019). Furthermore, we calculated the difference between the model prediction and the observed value. Standard errors or 95% confidence intervals were reported alongside each measurement to allow the reader for independent data review. To validate or deny any statistical anomalies in the number of deaths during 2020, we checked all the trends of the following annual statistics within the 2011-2019 time frame: male deaths by age group, female deaths by age group, male mortality by age group, female mortality by age group, deaths by causes of death, male deaths by causes of death, female deaths by causes of death. Specifically, we searched for anomalous non-linear sub-trends capable of distorting the interpretations on the cumulative data (indeed, sum of linear trends is linear as shown in Supplementary File 1, section “2. Linear trends”). An example of this phenomenon is shown in Figure 1. Concerning male and female deaths for age groups, we also calculated the 2020 forecast for each age group through an *ARIMA (p, d, q)* model using the *RStudio v*.*4*.*1*.*2* software (libraries *forecast* and *tseries*). The *d* parameter has been set equal to the number of differentiations necessary to sufficiently stationary the series. The series stationarity was evaluated through the Augmented Dickey-Fuller test, the Mann-Kendall test (to highlight possible trends), and a graphical control. The *p* and *q* parameters were chosen by examining autocorrelation graphs (ACF) and partial autocorrelation (PACF) graphs and then minimizing the Akaike Information Criteria (AIC) value. To facilitate the reproducibility of the analysis, we have provided all the ARIMA models in Supplementary File 2. Finally, we used OLS multiple linear regression to verify any correlations with demographic and geographic statistics such as population, population density, and latitude. The standard assumptions of the model (i.e., normality of the residuals, homoskedasticity, absence of outliers, and absence of multicollinearity) were automatically verified by the *Multiple Linear Regression Calculator* tool by *Statistics Kingdom* [22].

**Figure 1.**
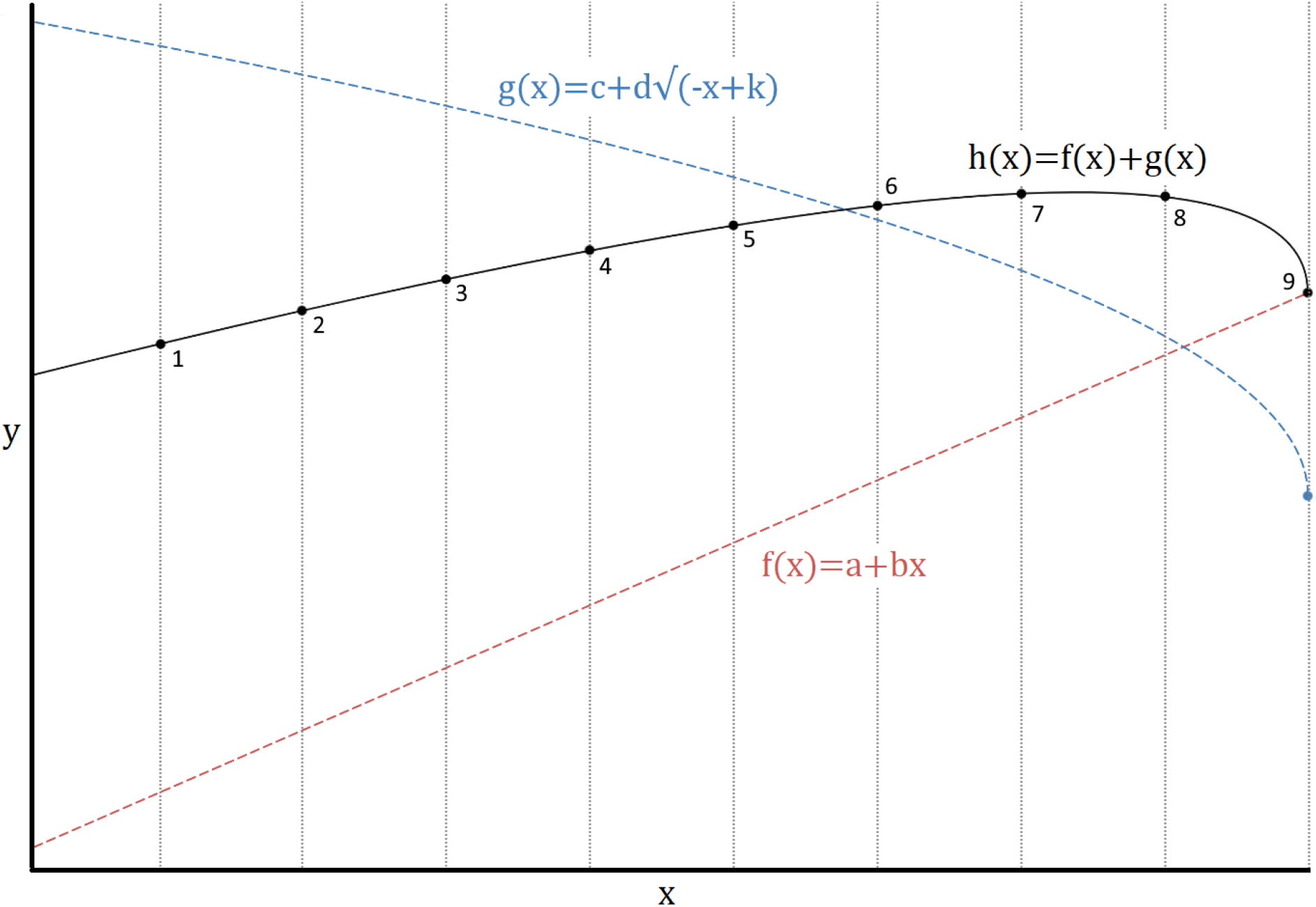
Example of a statistical anomaly due to the superposition of an increasing linear trend and a decreasing sublinear trend. Indeed, measure number 9 is very far from the prediction of a model built or trained on the first eight values. However, such an anomaly is not due to any external factor (ie, the reason is endogenous). Such a scenario could be even more complex to decipher if there are multiple trends to be considered.

## Results

### Overall deaths excess during 2020

Compared to the OLS linear regression model prediction (Figure 2), the 2020 excess in the observed number of deaths in Italy was significantly and substantially larger (*P=*.*003, EXC=89287, %EXC=13*.*6±5*.*3*). However, the significance level was not uniform across regions: indeed, the P-values of the northernmost regions - from Piedmont to Emilia Romagna - fluctuated between highly significant figures (*P<*.*001* or .001≤*P*≤.*011*). This area accounted for 50.4% of the whole excess deaths despite the theoretical prediction being 46.9%. After that, only Marche, Lazio, Puglia, and Sardinia reached modest P-values (*0*.*015*≤*P*≤*0*.*089*). The remaining regions recorded P-values of little significance (*P=*.*183* or *P>*.*300*). Nonetheless, it is worth mentioning that the number of deaths exceeded that predicted by the model in all Italian regions. The detailed report is presented in Tables S1, S2, and S3. Finally, Figure 2 also shows the high statistical confidence between the values predicted by the OLS linear regression and ARIMA (0,2,2) models; this constitutes further proof of the goodness of linear interpolation.

**Figure 2.**
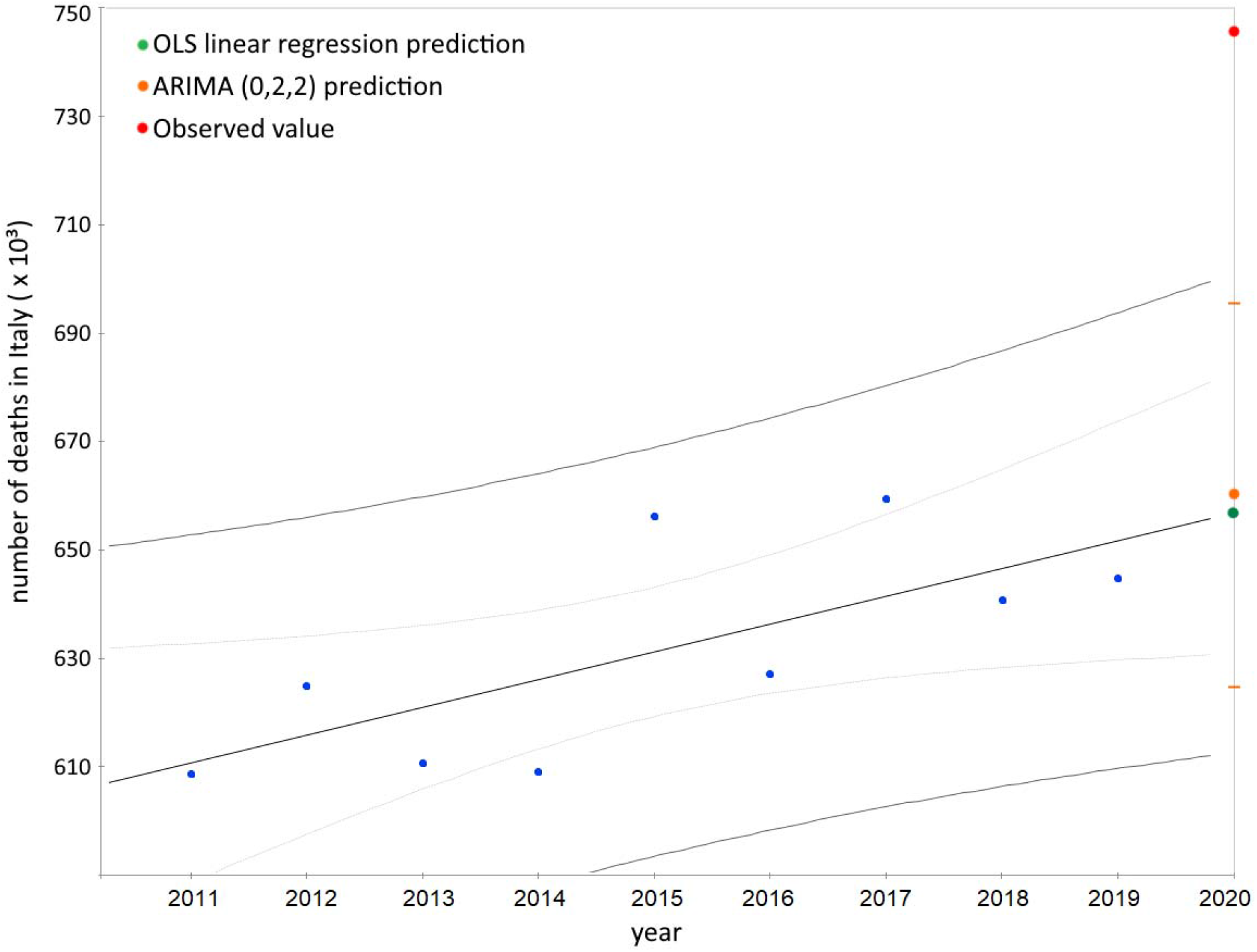
Excess of deaths in Italy during 2020: comparison between the prediction of the OLS linear regression and ARIMA (0,2,2) model with the observed value. The graph shows the number of annual deaths from 2011 to 2020 in Italy and the interpolating line. The narrow bands represent the linear regression 95% confidence interval of the mean value, while the wide bands represent the 95% confidence interval of the observed values from 2011 to 2019. The orange dashes represent the 95% confidence interval of the ARIMA prediction.

### Male mortality rate during 2020

When the male mortality rate is considered, the 2020 excesses were large and highly significant in 14/21 regions (*P<*.*001* or .*001*≤*P*≤.*013*). Moderate significant increases were observed in the other four regions (*0*.*042*≤*P*≤*0*.*077*). A low significance was obtained only in Molise, Basilicata, Calabria, and Sicily (*P>0*.*199*). Overall, the excess male mortality in Italy during 2020 was very high and markedly significant (*P<*.*001, EXC=18*.*8/10,000, %EXC=18*.*4±5*.*4*). Moreover, all regions recorded an increase in male mortality between 5% (Basilicata) and 38% (Lombardy). Details of each region are provided in Tables 1 and 2. Further information on the model goodness is provided in Table S4.

**Table 1.** Regional male mortality statistics: comparison of OLS linear regression model predictions and observed data. Legend: S.E. = standard error, PRED. = prediction.

| **REGION** | **PREDICTED** | **S.E.** | **OBSERVED** | **%** | **S.E.** | **%** |
| --- | --- | --- | --- | --- | --- | --- |
|  |  | **PRED.** |  | **EXCESS** | **EXCESS** |  |
| **Italy** | 102.1 | 4.6 | 120.9 | 18.4 | 5.4 |  |
| **Piemonte** | 105.7 | 5.2 | 132.3 | 25.1 | 6.2 |  |
| **Valle d'Aosta** | 114.4 | 11.3 | 136.3 | 19.1 | 12.3 |  |
| **Lombardia** | 98.5 | 4.1 | 136.2 | 38.3 | 5.8 |  |
| **Bolzano** | 93.6 | 4.9 | 110.0 | 17.5 | 6.2 |  |
| **Trento** | 90.7 | 4.7 | 121.0 | 33.4 | 7.0 |  |
| **Veneto** | 97.2 | 3.7 | 114.7 | 18.0 | 4.5 |  |
| **Friuli Venezia Giulia** | 99.6 | 5.9 | 116.3 | 16.8 | 7.0 |  |
| **Liguria** | 103.4 | 5.6 | 126.5 | 22.3 | 6.7 |  |
| **Emilia-Romagna** | 97.5 | 4.5 | 116.1 | 19.1 | 5.6 |  |
| **Toscana** | 97.2 | 6.0 | 108.5 | 11.6 | 7.0 |  |
| **Umbria** | 94.5 | 6.3 | 105.4 | 11.5 | 7.6 |  |
| **Marche** | 96.3 | 5.4 | 111.1 | 15.3 | 6.6 |  |
| **Lazio** | 100.1 | 5.2 | 110.1 | 10.0 | 5.7 |  |
| **Abruzzo** | 101.9 | 4.5 | 114.6 | 12.5 | 5.0 |  |
| **Molise** | 107.3 | 7.2 | 113.8 | 6.0 | 7.2 |  |
| **Campania** | 116.1 | 5.5 | 129.9 | 11.9 | 5.4 |  |
| **Puglia** | 100.1 | 5.8 | 115.6 | 15.5 | 6.7 |  |
| **Basilicata** | 107.2 | 6.1 | 112.9 | 5.3 | 6.0 |  |
| **Calabria** | 106.5 | 6.1 | 113.9 | 6.9 | 6.2 |  |
| **Sicilia** | 112.7 | 7.2 | 122.9 | 9.1 | 7.0 |  |
| **Sardegna** | 101.2 | 5.3 | 113.7 | 12.3 | 5.9 |  |

**Table 2.**
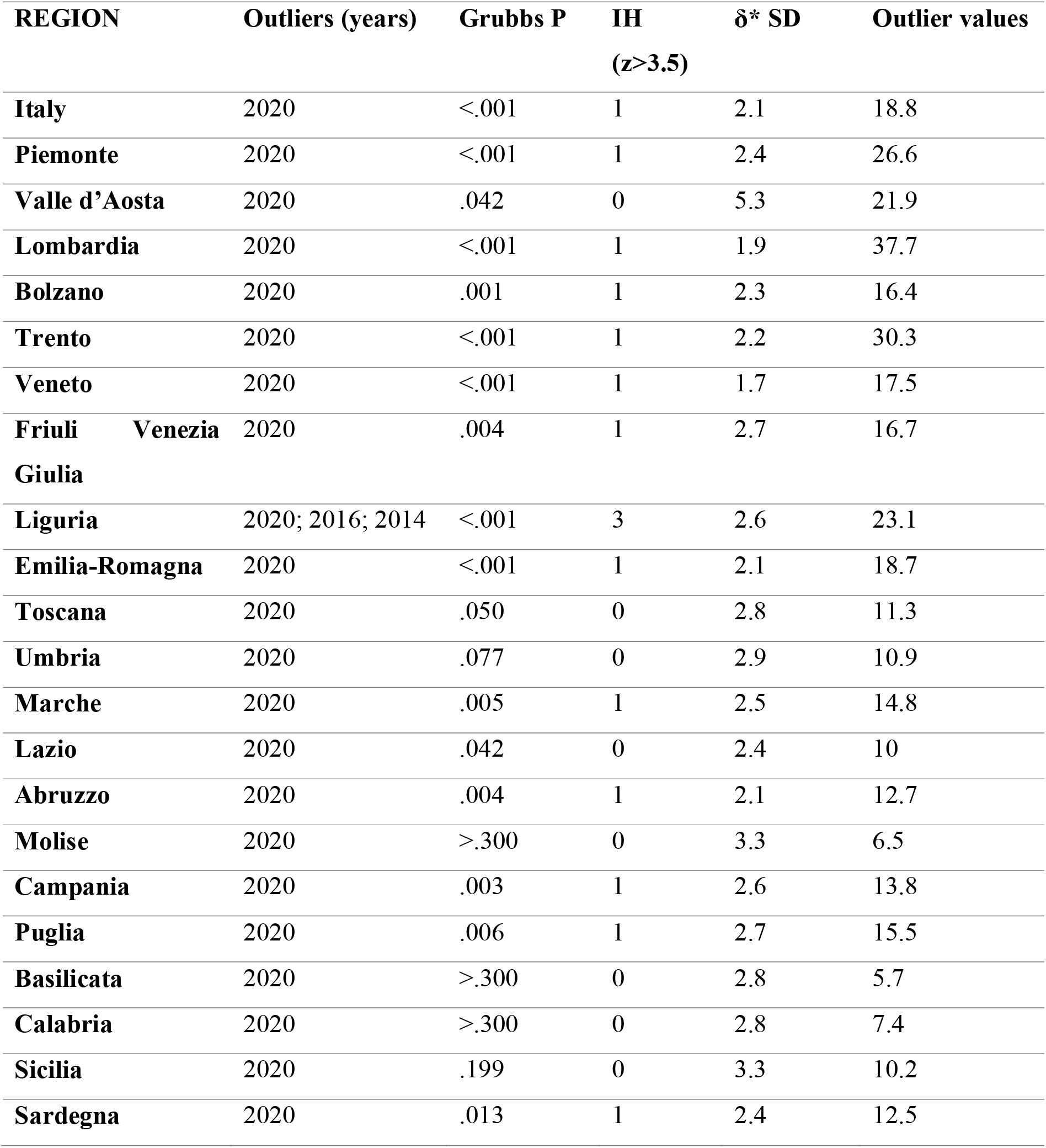
Statistical significance of excess male mortality rates during 2020: regional scenario. Legend: Grubbs P = Grubbs test P-value, IH = Iglewicz-Hoaglin test result, δ* SD = residuals standard deviation.

| REGION | Outliers (years) | Grubbs P | IH<br>(z>3.5) | $\delta^*$ SD | Outlier values |
| --- | --- | --- | --- | --- | --- |
| Italy | 2020 | <.001 | 1 | 2.1 | 18.8 |
| Piemonte | 2020 | <.001 | 1 | 2.4 | 26.6 |
| Valle d'Aosta | 2020 | .042 | 0 | 5.3 | 21.9 |
| Lombardia | 2020 | <.001 | 1 | 1.9 | 37.7 |
| Bolzano | 2020 | .001 | 1 | 2.3 | 16.4 |
| Trento | 2020 | <.001 | 1 | 2.2 | 30.3 |
| Veneto | 2020 | <.001 | 1 | 1.7 | 17.5 |
| Friuli Venezia Giulia | 2020 | .004 | 1 | 2.7 | 16.7 |
| Liguria | 2020; 2016; 2014 | <.001 | 3 | 2.6 | 23.1 |
| Emilia-Romagna | 2020 | <.001 | 1 | 2.1 | 18.7 |
| Toscana | 2020 | .050 | 0 | 2.8 | 11.3 |
| Umbria | 2020 | .077 | 0 | 2.9 | 10.9 |
| Marche | 2020 | .005 | 1 | 2.5 | 14.8 |
| Lazio | 2020 | .042 | 0 | 2.4 | 10 |
| Abruzzo | 2020 | .004 | 1 | 2.1 | 12.7 |
| Molise | 2020 | >.300 | 0 | 3.3 | 6.5 |
| Campania | 2020 | .003 | 1 | 2.6 | 13.8 |
| Puglia | 2020 | .006 | 1 | 2.7 | 15.5 |
| Basilicata | 2020 | >.300 | 0 | 2.8 | 5.7 |
| Calabria | 2020 | >.300 | 0 | 2.8 | 7.4 |
| Sicilia | 2020 | .199 | 0 | 3.3 | 10.2 |
| Sardegna | 2020 | .013 | 1 | 2.4 | 12.5 |

### Female mortality rate during 2020

As for the increase in female mortality during 2020, the scenario was more similar to that of overall deaths. Indeed, highly significant increases were found in the northern regions and Sardinia (*P<*.*001* or .*001*≤*P*≤.*019*, except Valle d’Aosta, *P=*.*042*). Moderately and scarcely significant increases were recorded in the rest of Italy (*P>*.*098*). Nevertheless, all regions experienced an increase in female mortality rate between 4% (Basilicata) and 31% (Lombardy). Details of each region are provided in Tables 3 and 4. Further information on the model goodness is provided in Table S5.

**Table 3.**
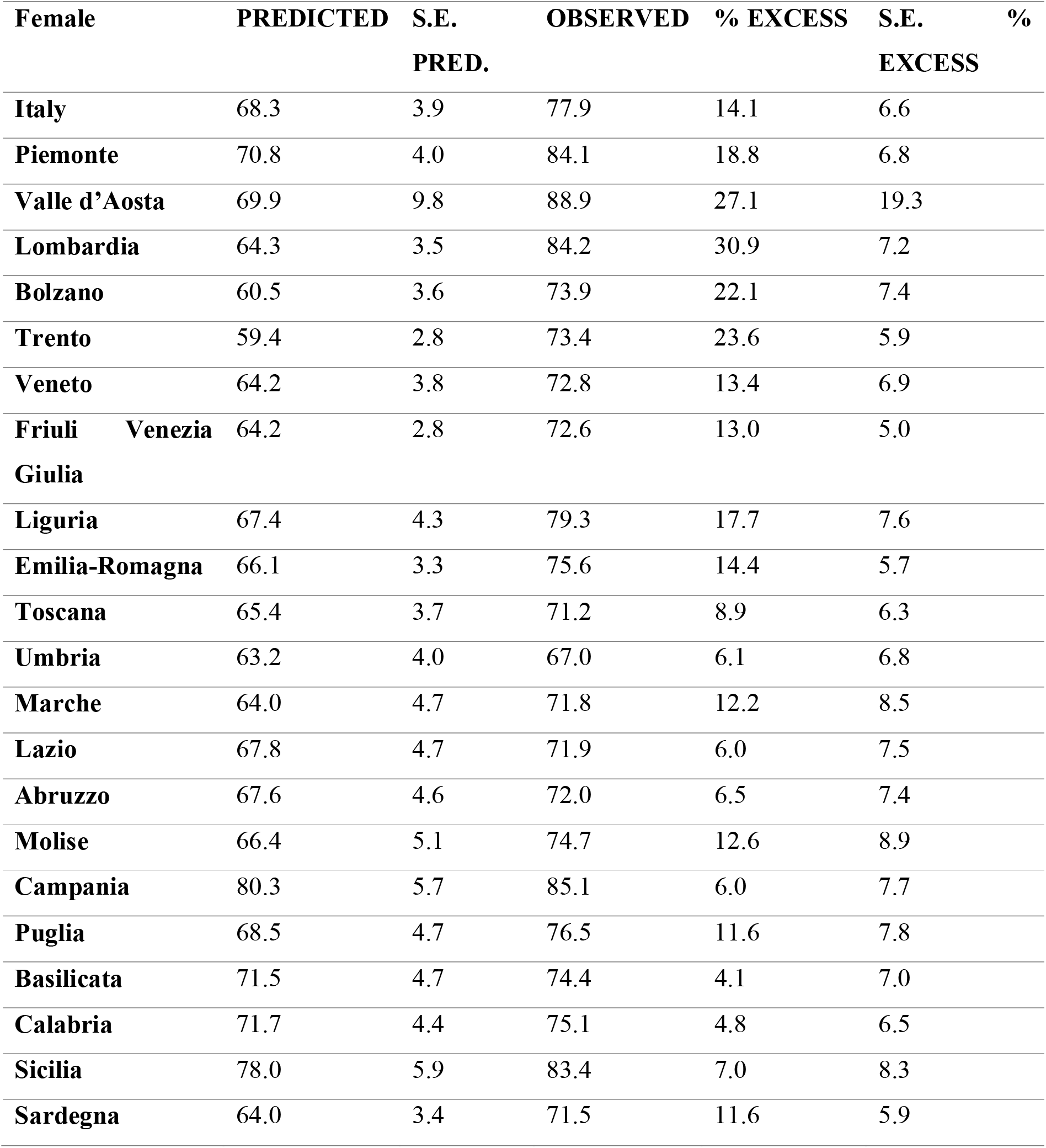
Regional female mortality statistics: comparison of OLS linear regression model predictions and observed data. Legend: S.E. = standard error, PRED. = prediction.

| **Female** | **PREDICTED** | **S.E.** | **OBSERVED** | **% EXCESS** | **S.E.** | **%** |
| --- | --- | --- | --- | --- | --- | --- |
|  |  | **PRED.** |  |  | **EXCESS** |  |
| **Italy** | 68.3 | 3.9 | 77.9 | 14.1 | 6.6 |  |
| **Piemonte** | 70.8 | 4.0 | 84.1 | 18.8 | 6.8 |  |
| **Valle d’Aosta** | 69.9 | 9.8 | 88.9 | 27.1 | 19.3 |  |
| **Lombardia** | 64.3 | 3.5 | 84.2 | 30.9 | 7.2 |  |
| **Bolzano** | 60.5 | 3.6 | 73.9 | 22.1 | 7.4 |  |
| **Trento** | 59.4 | 2.8 | 73.4 | 23.6 | 5.9 |  |
| **Veneto** | 64.2 | 3.8 | 72.8 | 13.4 | 6.9 |  |
| **Friuli Venezia Giulia** | 64.2 | 2.8 | 72.6 | 13.0 | 5.0 |  |
| **Liguria** | 67.4 | 4.3 | 79.3 | 17.7 | 7.6 |  |
| **Emilia-Romagna** | 66.1 | 3.3 | 75.6 | 14.4 | 5.7 |  |
| **Toscana** | 65.4 | 3.7 | 71.2 | 8.9 | 6.3 |  |
| **Umbria** | 63.2 | 4.0 | 67.0 | 6.1 | 6.8 |  |
| **Marche** | 64.0 | 4.7 | 71.8 | 12.2 | 8.5 |  |
| **Lazio** | 67.8 | 4.7 | 71.9 | 6.0 | 7.5 |  |
| **Abruzzo** | 67.6 | 4.6 | 72.0 | 6.5 | 7.4 |  |
| **Molise** | 66.4 | 5.1 | 74.7 | 12.6 | 8.9 |  |
| **Campania** | 80.3 | 5.7 | 85.1 | 6.0 | 7.7 |  |
| **Puglia** | 68.5 | 4.7 | 76.5 | 11.6 | 7.8 |  |
| **Basilicata** | 71.5 | 4.7 | 74.4 | 4.1 | 7.0 |  |
| **Calabria** | 71.7 | 4.4 | 75.1 | 4.8 | 6.5 |  |
| **Sicilia** | 78.0 | 5.9 | 83.4 | 7.0 | 8.3 |  |
| **Sardegna** | 64.0 | 3.4 | 71.5 | 11.6 | 5.9 |  |

**Table 4.** Statistical significance of excess female mortality rates during 2020: regional scenario. Legend: Grubbs P = Grubbs test P-value, IH = Iglewicz-Hoaglin test result, δ* SD = residuals standard deviation.

|  | Outliers (years) | Grubbs P | IH<br>(z>3.5) | δ* SD | Outlier<br>values |
| --- | --- | --- | --- | --- | --- |
| **Italy** | 2020 | .010 | 1 | 1.8 | 9.6 |
| **Piemonte** | 2020 | .001 | 1 | 1.9 | 13.3 |
| **Valle d'Aosta** | 2020 | .042 | 1 | 4.5 | 19 |
| **Lombardia** | 2020 | <.001 | 1 | 1.6 | 19.9 |
| **Bolzano** | 2020 | <.001 | 1 | 1.7 | 13.4 |
| **Trento** | 2020 | <.001 | 1 | 1.3 | 14 |
| **Veneto** | 2020 | .019 | 1 | 1.8 | 8.6 |
| **Friuli Venezia Giulia** | 2020 | .003 | 1 | 1.3 | 8.4 |
| **Liguria** | 2020 | .005 | 1 | 2 | 11.9 |
| **Emilia-Romagna** | 2020 | .003 | 1 | 1.5 | 9.5 |
| **Toscana** | 2020 | .133 | 0 | 1.7 | 5.8 |
| **Umbria** | 2020 | >.300 | 0 | 1.8 | 3.8 |
| **Marche** | 2020; 2017 | .098 | 2 | 2.2 | 7.8 |
| **Lazio** | 2014 | >.300 | 0 | 2.2 | 4.1 |
| **Abruzzo** | 2020 | >.300 | 0 | 2.1 | 4.4 |
| **Molise** | 2020 | .113 | 0 | 2.4 | 8.3 |
| **Campania** | 2015 | >.300 | 0 | 2.7 | 4.8 |
| **Puglia** | 2020 | .086 | 0 | 2.2 | 8 |
| **Basilicata** | 2015 | >.300 | 0 | 2.2 | 2.9 |
| **Calabria** | 2020 | >.300 | 0 | 2 | 3.4 |
| **Sicilia** | 2020 | >.300 | 0 | 2.8 | 5.5 |
| **Sardegna** | 2020 | .017 | 1 | 1.6 | 7.5 |

### Relationship between deaths and geographical-demographic statistics

The linear multi-regression model among the log-transformed statistics the *regional number of inhabitants* (*X*_*1*_), *regional population density* (*X*_*2*_), *regional latitude* (*X*_*3*_), and *2020 regional excess deaths* (*Y*), returned the following equation: *Y=f(X*_*3*_*)=k×pow(X*_*3*_, *a)*, with *R=0*.*82, p<*.*001*. Therefore, the excess of deaths is more correlated with latitude than with the other two demographic statistics (*R*_*1,2*_ *=-*.*06, R*_*1,3*_ *=*.*38*).

### Retrospective analysis of deaths

Figure 3 shows the number of deaths per cause of death from 2012 to 2017 in Italy. We can observe that all-time series are linear growing or stationary. This provides strong evidence against the hypothesis of anomalous non-linear sub-trends (ASTs) at the national level. Furthermore, Figure 3 shows a large gap between the first two causes of death and the other seven: indeed, tumors and diseases of the circulatory system always accounted for over 60% of total deaths (also considering the projections for 2020). The male (female) percentages of deaths for tumors ranged from 55.6% (44.4%) to 56.3% (43.7%), while deaths related to the circulatory system were 43.1% (56.9%) to 43.7% (56.3%). Such a result rules out sex-related ASTs for these two classes of pathologies.

**Figure 3.**
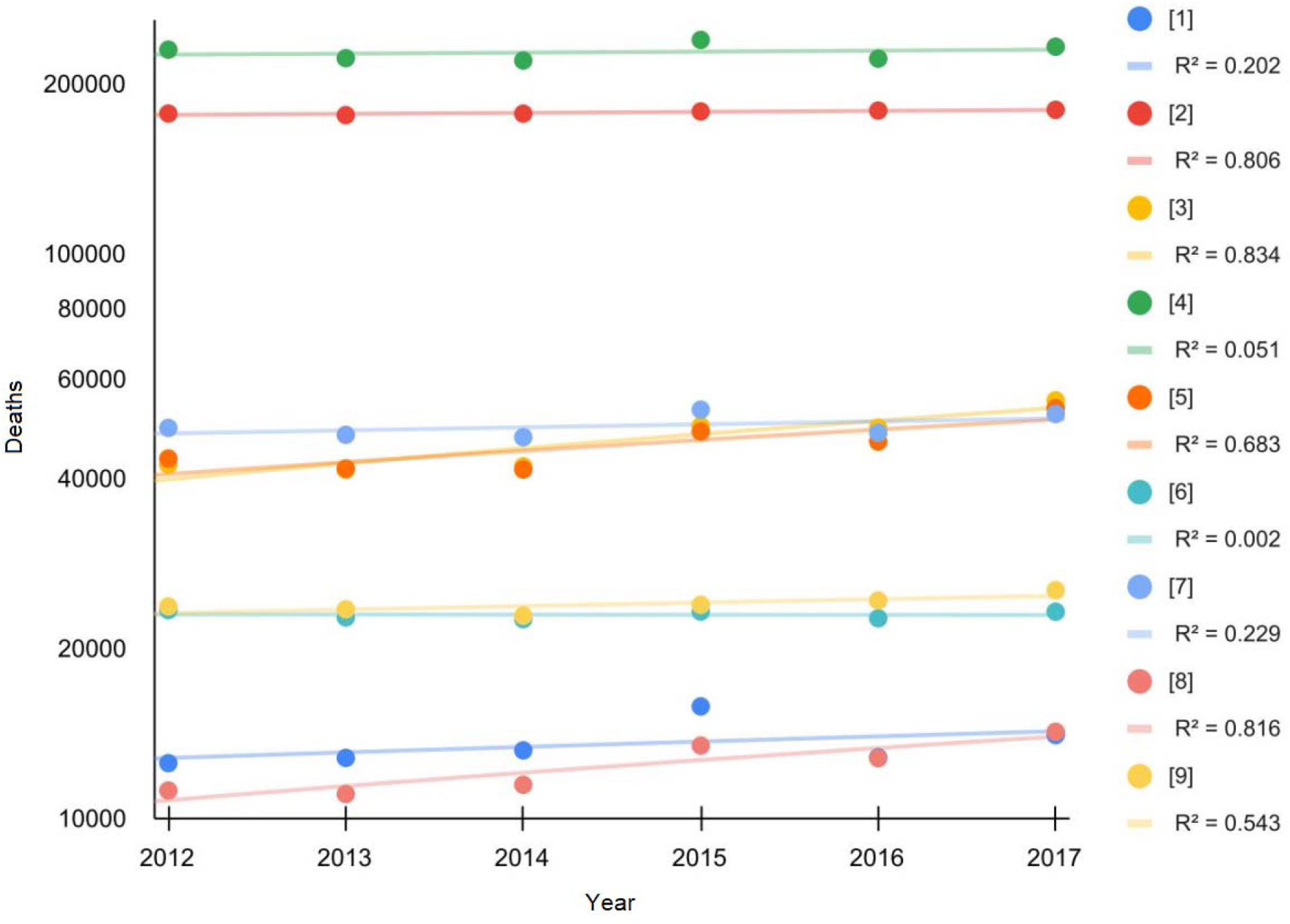
Number of deaths per cause of death from 2011 to 2017 in Italy. Legend: [1] = Infectious and parasitic diseases, [2] = Tumors, [3] = Psychic disorders, diseases of the nervous system and organs of the senses, [4] = Diseases of the circulatory system, [5] = Diseases of the respiratory system, [6] = Diseases of the digestive system, [7] = Other morbid states, [8] = Poorly defined symptoms, signs, and morbid states, [9] = External causes of trauma and poisoning.

Since also the male (female) percentage of total deaths from 2011 to 2019 fluctuated between 47.6% (52.4%) and 48.3% (51.7%), we can exclude any sex-related ASTs. Finally, Figure 4 and Figure 5 show male and female deaths by age group from 2011 to 2019. In particular, it is possible to observe the regularity of the patterns by age group. Similar results are presented in Figures S1 and S2, which show the mortality of each age group over the same time frame. Explicitly calculating each trend for each age and sex group and summing the predictions for 2020, we obtained the best value of 648,733 deaths (fully compatible with the prediction of the OLS linear regression and ARIMA models, i.e., 656,859 and 659,547 deaths). All trends were markedly linear (Figure S3 and S4). Summing up all the forecasts of the ARIMA models for each age and sex group, we obtained a total of 637,534 deaths, which is even lower than previous predictions. A similar result was obtained by summing the global trends of males and females (640,508 deaths). These data confirm the absence of sex-related ASTs and provide decisive evidence on the absence of age-related ASTs. Therefore, this analysis provides strong evidence in favor of a marked and significant level-shift in the time-series representing the annual deaths in Italy from 2011 to 2020, and the absence of hidden anomalous deaths and mortality trends capable of justifying the above level-shift.

**Figure 4.**
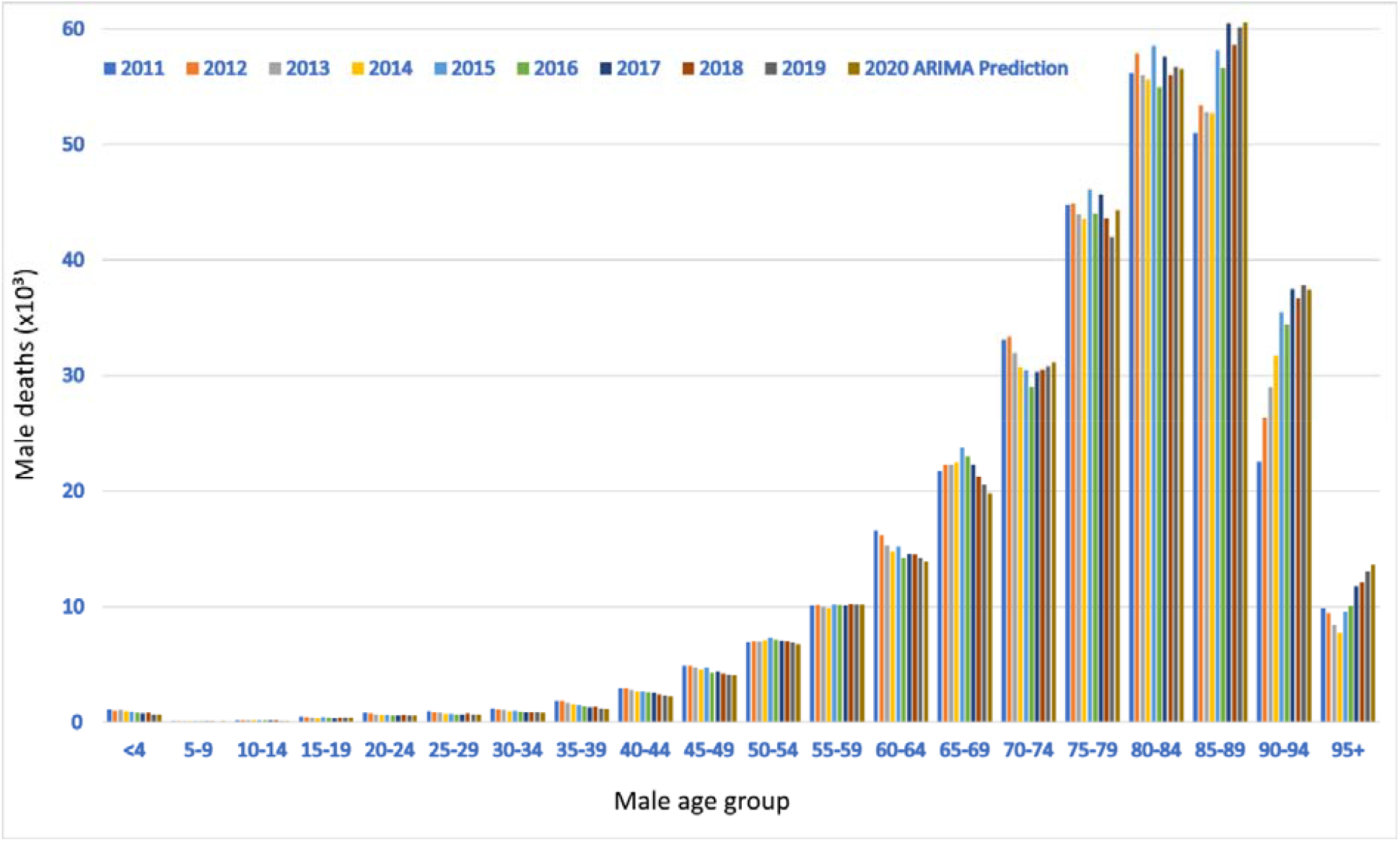
Male deaths per age group from 2011 to 2019 and ARIMA predictions for 2020.

**Figure 5.**
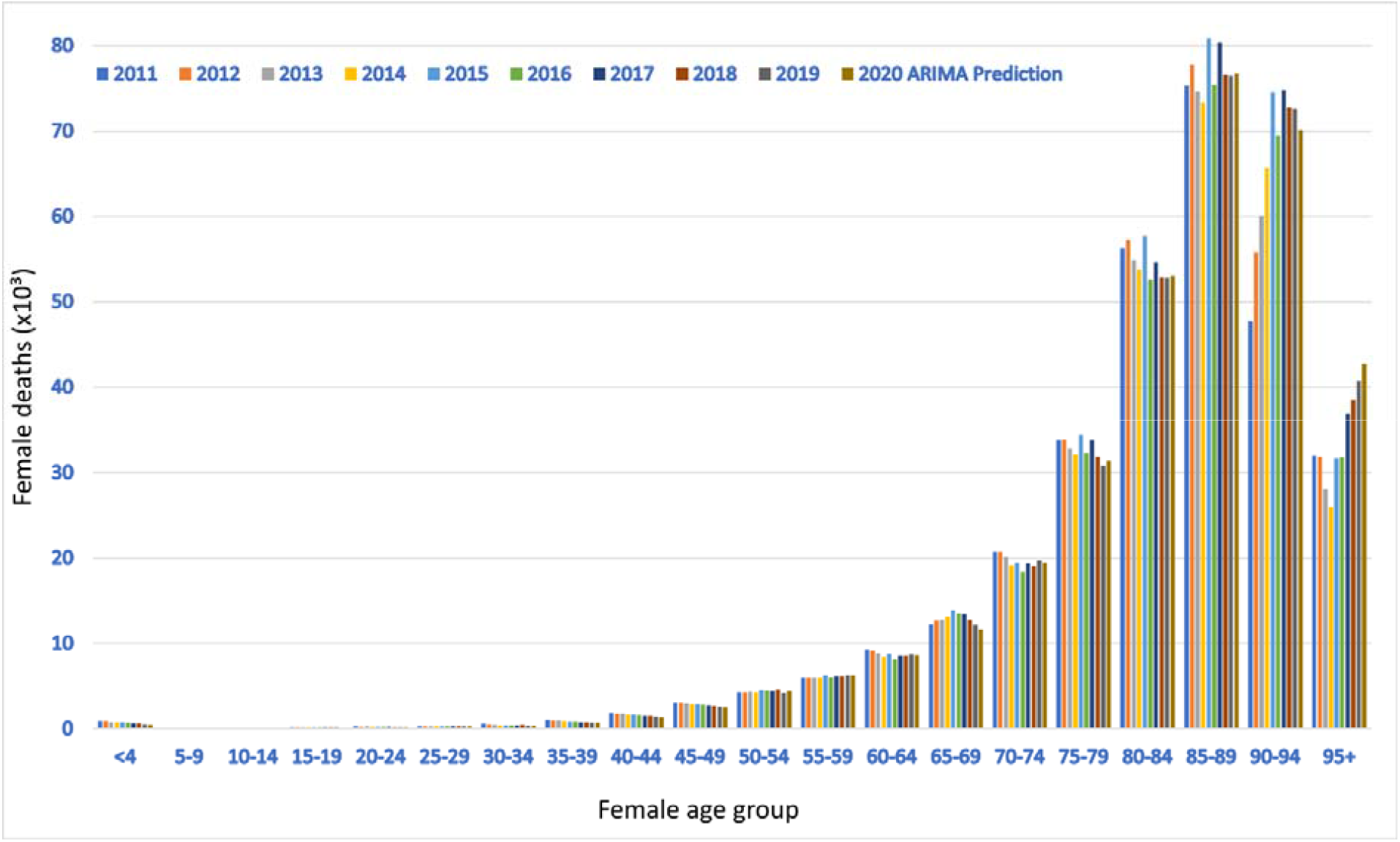
Female deaths per age group from 2011 to 2019 and ARIMA predictions for 2020.

## Discussion

### Main findings

This paper provides strong evidence in favor of an anomalous mortality event during 2020 in Italy, which was not predictable based on endogenous causes such as deaths and mortality trends between 2011 and 2019. Notably, the number of total deaths observed in 2020 exceeded the linear regression model prediction by more than 89,000 (a value nearly three times greater than the prediction standard error) and the ARIMA prediction by more than 86,000. Grubbs and Iglewicz-Hoaglin tests confirmed that this figure was unexpected. We also highlighted marked gender differences: indeed, total excess male mortality (*+18%*) was substantially higher than total excess female mortality (*+14%*). The total excess mortality was positively correlated with latitude, which explained the dataset variability much better than demographic statistics like population number and density. All the “deceases due to causes of death” trends from 2012 to 2017 were appreciably linear or stationary; this precludes the existence of anomalous sub-trends linked to the causes of death. Moreover, summing up all the 2020 deaths predictions by age group, we obtained a value ranging from 640,000 to 660,000 deceases, significantly far from the observed one (750,000). In conclusion, these findings confirm the absence of any confounding inner sub-trends capable of explaining the excess deaths during 2020 in Italy.

To the best of our knowledge, the most comprehensive and detailed study examining the excess mortality during 2020 in Italy was the report redacted by the National Institute of Statistics (ISTAT) and National Institute of Health (ISS) [23]. Their research focuses on comparing the March-December 2015-2019 and 2020 periods starting from the assumption that COVID-19 is the cause of the discrepancies observed. On the contrary, our analysis has been more impartial since we have not introduced any hypothesis about the reasons that caused this phenomenon. Therefore, our findings provide evidence of statistical and epidemiological significance that had not been considered before. Specifically, excluding internal causes gives further strength to the theories that identify COVID-19 as the principal cause of such a tragic scenario. COVID-19 dangerousness is confirmed at the molecular-genetic level [24]: compared to its predecessor, SARS-CoV-2 binds to the ACE2 peptidase domain two-four times more strongly, which increases its severity [25–27]. Besides, our results statistically support the greater virulence and mortality of COVID-19 in northern Italy and towards the male population [28–30]. As of November 2021, there are numerous well-targeted hypotheses, epidemiological models, and clinical evidence capable of justifying these asymmetries. For example, on average, Italian men drink and smoke significantly more than Italian women and are less likely to comply with anti-COVID-19 regulations [31, 32, 33]. This exposes them to considerably greater risks of critical COVID-19 outcomes [34–37]. Alongside this, various studies provide complementary explanations based on sexual dimorphisms [38]. Some in vivo experiments have shown the possible role of sex hormones in regulating ACE2 activity, essential for SARS-CoV-2 to enter the host cells in upper and lower respiratory tracts [39]. Regarding geographic differences, an increasing number of mathematical-statistical investigations classify COVID-19 as a seasonal low-temperature infection [40, 41], although the effect size of the environmental factors is still debated [42]. However, it is a fact that low temperatures can have indirect effects on the spread of infections, like the creation of indoor gatherings - with insufficient air circulation - and the weakening of the immune defenses [45–44]. Since average temperatures in northern regions are lower than the rest of the peninsula [46], this phenomenon could partially explain the Italian epidemiological scenario. A large literature has also identified pollution as a relevant COVID-19 risk factor. For instance, NO2, PM10, and PM2.5 were causally connected with more serious situations, as they can drastically reduce the immune response and compromise respiratory functions [46–48]. This type of pollutant is widespread in the Po Valley [46, 47]. Similar conclusions were achieved by Pluchino et al., who developed a novel model for epidemic risk assessment in Italy [49]. Nonetheless, the impact of COVID-19 is influenced by numerous comorbidities, such as cancer, chronic kidney diseases, diabetes mellitus, hypertension, chronic obstructive pulmonary diseases, asthma, chronic respiratory diseases, immunocompromised state, HIV infection, heart conditions, overweight and obesity, dementia or other neurological conditions, and mental health conditions [50–52]. The majority of these pathologies are more common in the older age groups, which helps explain the greater aggressiveness of the infection in some regions [17, 49]. Hence, it is necessary to consider that the pre-pandemic epidemiological scenario has contributed to enhancing the disease damage in Italy. Nevertheless, it would be incorrect to consider only the older population as vulnerable: phenomena such as long COVID (i.e., the onset of medical complications that last weeks to months after initial recovery) are increasing in younger age groups, including children and adolescents [53, 54]. The most common symptoms of long COVID are fatigue, weakness, cough, chest tightness, breathlessness, palpitations, myalgia, and difficulty focusing; their appearance is not related to the severity of the COVID-19 course [54, 55]. Moreover, new variants of concern (VOCs) - favored by the uncontrolled spread of the virus - continuously pose new threats to all age groups [24, 56, 57]. To date, the effectiveness of vaccination against the most hostile variants such as Delta decreased but remained sufficiently high (approximately 60-90% depending on the vaccine used) [58]. For example, the BNT162b2 vaccine (two doses) maintained an efficacy of around 90% against 60% of ChAdOx1 (two doses) [59]. These data confirm vaccines as a fundamental tool to combat the spread of the pandemic. BNT162b2 is also effective in reducing viral loads of breakthrough infections, thus decreasing the chances of infecting others even in the rare cases where the disease is contracted. Overall, mRNA vaccines conferred a lesser risk of SARS-COV-2 infection compared to adenoviral vector vaccines [60]. Nonetheless, heterologous vaccination (ie, ChAdOx1 as first dose and BNT162b2 as second dose) showed stronger neutralizing activity against VOCs [61]. COVID-19 vaccines have also decreased hospitalizations, intensive care admissions, and deaths by over 90% in the vaccinated Italian population [62]. This allows countries to maintain performing health services, fundamental to delay the peaks of any future epidemiological curves and thus diminish the number of deaths [63]. The safety of COVID-19 vaccines is constantly monitored: the side effects encountered so far are very rare (much more so than other commonly used drugs) [64]. However, any vaccination strategy cannot replace other non-pharmaceutical interventions, which remain essential in controlling and isolating new outbreaks [65]. While this paper has provided evidence in favor of a high number of deaths due to COVID-19 in Italy, containment measures such as lockdowns, social distancing and masks have prevented the death toll from being numerous times higher [66–69].

### Practical implications

Our analysis confers statistical significance to the discoveries of previous and literature, providing strong evidence against potential endogenous causes for the mortality increase recorded in Italy during 2020. Combining our findings with the most recent scientific evidence (discussed in the previous subsection), we can state that the impact of COVID-19 on excess deaths during 2020 in Italy constitutes a scientific fact. This answers the question, “Died from COVID or died with COVID?”. Specifically, this manuscript can be exploited by health authorities and disclosure agencies to discredit fake news that minimizes the COVID-19 risk. Moreover, given the significant concordance between the predictions of the ARIMA and OLS regression models, we suggest that these models be exploited to predict mortality trends.

## Data Availability

All data produced in the present work are contained in the manuscript

