## Supplementary file 1 for "Dying From COVID-19 or With COVID-19: A Definitive Answer Through a Retrospective Analysis of Mortality in Italy"

**1. Data collection details**

[Number of deaths by age group]. Each age group has been manually selected from the website: <http://dati.istat.it/Index.aspx?DataSetCode=DCIS_MORTALITA1#> (item: mortalità - decessi - morti). The datasets have been divided into male, female, and total. Investigated period: 2011-2019. We have left all the remaining parameters to the default ones.

[Number of deaths]. We used the following Excel file already paginated by the national health observatory: <https://www.osservatoriosullasalute.it/wp-content/uploads/2021/05/ro-2020-isc-covid.xlsx>. The datasets have been divided into male, female, and total. Investigated period: 2011-2020.

[Number of deaths by death causes]. The data were manually extracted from item "Morti per classe di età, sesso e gruppo di cause" of the following reports of the National Statistical Institute (ISTAT): <https://www.istat.it/it/files//2020/12/C04.pdf>, <https://www.istat.it/it/files//2019/12/C04.pdf>, <https://www.istat.it/it/files//2018/12/C04.pdf>, <https://www.istat.it/it/files//2017/12/C04.pdf>, <https://www.istat.it/it/files//2016/12/C04.pdf>, <https://www.istat.it/it/files//2015/12/C04.pdf>. The datasets have been divided into male, female, and total. Investigated period: 2012-2017.

[Population number per age group]. The data were manually extracted from: <https://www.tuttitalia.it/statistiche/popolazione-eta-sesso-stato-civile-2019/>. The datasets have been divided into male, female, and total. Investigated period: 2011-2019.

[Population number and density per region]. The data were manually extracted from: <https://www.tuttitalia.it/regioni/densita/>. Investigated period: 2020.

All websites were consulted on November 01, 2021.

**2. Linear trends**

f(x) = ax+b, g(x) = cx+d | f(x)+g(x) = ax+b+cx+d = (a+c)x+(b+d) = ex+f.

**3. Supplementary Figures and Tables.**

|  | PRE | S.E. PRE | OBS | % EXC | S.E. % EXC |
| --- | --- | --- | --- | --- | --- |
| Italy | 656859 | 30563 | 746146 | 13.6 | 5.3 |
| Piemonte | 55070 | 2600 | 66054 | 19.9 | 5.7 |
| Valle d’Aosta | 1523 | 135 | 1849 | 21.4 | 11.1 |
| Lombardia | 102865 | 4165 | 136249 | 32.5 | 5.4 |
| Bolzano | 4590 | 175 | 5458 | 18.9 | 4.6 |
| Trento | 5246 | 167 | 6626 | 26.3 | 4 |
| Veneto | 50680 | 2077 | 57836 | 14.1 | 4.7 |
| Friuli Venezia Giulia | 14665 | 659 | 16617 | 13.3 | 5.1 |
| Liguria | 22023 | 1189 | 25827 | 17.3 | 6.4 |
| Emilia-Romagna | 51508 | 2309 | 59665 | 15.8 | 5.2 |
| Toscana | 44603 | 2445 | 48135 | 7.9 | 6 |
| Umbria | 10545 | 654 | 11131 | 5.6 | 6.6 |
| Marche | 18061 | 1159 | 20123 | 11.4 | 7.3 |
| Lazio | 60036 | 2952 | 62161 | 3.5 | 5.1 |
| Abruzzo | 15375 | 822 | 16296 | 6 | 5.7 |
| Molise | 3865 | 264 | 4127 | 6.8 | 7.4 |
| Campania | 56164 | 3122 | 59425 | 5.8 | 6 |
| Puglia | 40770 | 2312 | 44650 | 9.5 | 6.3 |
| Basilicata | 6654 | 284 | 6839 | 2.8 | 4.4 |
| Calabria | 20837 | 1125 | 21331 | 2.4 | 5.6 |
| Sicilia | 54351 | 3238 | 56753 | 4.4 | 6.3 |
| Sardegna | 17427 | 676 | 18994 | 9 | 4.3 |

**Supplementary Table 1.** Comparison between prediction and the actual number of deaths during 2020 in all Italian regions. Legend: PRE = predicted value, S.E. = standard error, OBS = observed value, % EXC = percentage excess

|  | OUT Period | Grubbs P | IH (z>3.5) | δ* SD | 2020 Exc |
| --- | --- | --- | --- | --- | --- |
| Italy | 2020 | 0.003 | 1 | 14171 | 89287 |
| Piemonte | 2020 | <.001 | 1 | 1206 | 10984 |
| Valle d’Aosta | 2020 | 0.011 | 1 | 63 | 326 |
| Lombardia | 2020 | <.001 | 1 | 1931 | 33384 |
| Bolzano | 2020 | <.001 | 1 | 81 | 868 |
| Trento | 2020 | <.001 | 1 | 78 | 1380 |
| Veneto | 2020 | 0.001 | 1 | 963 | 7156 |
| Friuli Venezia Giulia | 2020 | 0.003 | 1 | 305 | 1952 |
| Liguria | 2020 | 0.003 | 1 | 551 | 3804 |
| Emilia-Romagna | 2020 | <.001 | 1 | 1071 | 8157 |
| Toscana | 2020 | 0.183 | 0 | 1134 | 3532 |
| Umbria | 2020 | >0.3 | 1 | 303 | 586 |
| Marche | 2020 | 0.066 | 1 | 537 | 2062 |
| Lazio | 2020 | <.001 | 0 | 1369 | 2125 |
| Abruzzo | 2020 | >0.3 | 0 | 381 | 921 |
| Molise | 2020 | >0.3 | 0 | 122 | 262 |
| Campania | 2020; 2015 | >.03 | 2 | 1447 | 3261 |
| Puglia | 2020 | 0.089 | 1 | 1072 | 3880 |
| Basilicata | 2015 | >0.3 | 0 | 132 | 185 |
| Calabria | 2017 | >0.3 | 0 | 522 | 494 |
| Sicilia | 2015 | >0.3 | 0 | 1501 | 2402 |
| Sardegna | 2020 | 0.015 | 1 | 314 | 1567 |

**Supplementary Table 2.** Statistical significance of the excess deaths during 2020. Legend: OUT Period = outlier period, Grubbs P = Grubbs test P-value, IH = Iglewicz-Hoaglin test result, δ* SD = residuals standard deviation, 2020 Exc = 2020 deaths excess.

|  | F P | R² | S.E. R² | Δ* SW P | Δ SW P |
| --- | --- | --- | --- | --- | --- |
| Italy | 0.034 | 0.495 | 0.19 | 0.274 | 0.005 |
| Piemonte | 0.012 | 0.615 | 0.161 | 0.867 | <.001 |
| Valle d’Aosta | 0.043 | 0.464 | 0.195 | 0.178 | 0.009 |
| Lombardia | 0.003 | 0.744 | 0.118 | 0.908 | <.001 |
| Bolzano | 0.002 | 0.765 | 0.152 | 0.165 | <.001 |
| Trento | 0.001 | 0.795 | 0.098 | 0.74 | <.001 |
| Veneto | 0.009 | 0.645 | 0.152 | 0.718 | 0.003 |
| Friuli Venezia Giulia | 0.933 | 0.001 | 0.177 | 0.778 | 0.009 |
| Liguria | 0.883 | 0.003 | 0.029 | 0.709 | 0.005 |
| Emilia-Romagna | 0.069 | 0.398 | 0.203 | 0.627 | 0.002 |
| Toscana | 0.228 | 0.2 | 0.191 | 0.325 | 0.192 |
| Umbria | 0.514 | 0.063 | 0.126 | 0.249 | 0.205 |
| Marche | 0.16 | 0.261 | 0.202 | 0.052 | 0.031 |
| Lazio | 0.035 | 0.494 | 0.19 | 0.02 | 0.016 |
| Abruzzo | 0.428 | 0.092 | 0.147 | 0.042 | 0.049 |
| Molise | 0.954 | 0.001 | 0.017 | 0.025 | 0.039 |
| Campania | 0.11 | 0.323 | 0.206 | 0.023 | 0.022 |
| Puglia | 0.029 | 0.519 | 0.185 | 0.268 | 0.129 |
| Basilicata | 0.008 | 0.658 | 0.148 | 0.616 | 0.699 |
| Calabria | 0.091 | 0.355 | 0.205 | 0.12 | 0.223 |
| Sicilia | 0.149 | 0.273 | 0.203 | 0.478 | 0.297 |
| Sardegna | 0.001 | 0.796 | 0.097 | 0.817 | 0.052 |

**Supplementary Table 3.** Goodness-of-fit statistics for total excess deaths during 2020. Legend: F P = F-statistic P-value, S.E. = standard error, SW P = Shapiro-WIlk P-value.

|  | F P | R² | S.E. R² | Δ* SW P | Δ SW P |
| --- | --- | --- | --- | --- | --- |
| Italy | <.001 | 0.866 | 0.067 | 0.799 | 0.001 |
| Piemonte | 0.001 | 0.798 | 0.096 | 0.819 | <.001 |
| Valle d’Aosta | 0.28 | 0.164 | 0.181 | 0.334 | 0.084 |
| Lombardia | <.001 | 0.897 | 0.052 | 0.717 | <.001 |
| Bolzano | <.001 | 0.844 | 0.077 | 0.077 | 0.001 |
| Trento | <.001 | 0.887 | 0.057 | 0.403 | <.001 |
| Veneto | <.001 | 0.924 | 0.039 | 0.636 | 0.003 |
| Friuli Venezia Giulia | <.001 | 0.888 | 0.056 | 0.379 | 0.003 |
| Liguria | 0.001 | 0.817 | 0.088 | 0.014 | <.001 |
| Emilia-Romagna | 0.001 | 0.824 | 0.085 | 0.295 | <.001 |
| Toscana | 0.002 | 0.776 | 0.105 | 0.954 | 0.22 |
| Umbria | 0.001 | 0.823 | 0.086 | 0.349 | 0.214 |
| Marche | 0.002 | 0.782 | 0.103 | 0.401 | 0.007 |
| Lazio | <.001 | 0.867 | 0.066 | 0.975 | 0.244 |
| Abruzzo | <.001 | 0.903 | 0.049 | 0.08 | 0.004 |
| Molise | 0.002 | 0.753 | 0.115 | 0.213 | 0.198 |
| Campania | <.001 | 0.85 | 0.074 | 0.313 | 0.005 |
| Puglia | 0.001 | 0.804 | 0.094 | 0.299 | 0.009 |
| Basilicata | 0.01 | 0.636 | 0.155 | 0.967 | 0.864 |
| Calabria | 0.005 | 0.7 | 0.134 | 0.248 | 0.39 |
| Sicilia | 0.007 | 0.674 | 0.143 | 0.5 | 0.298 |
| Sardegna | 0.001 | 0.822 | 0.086 | 0.726 | 0.058 |

**Supplementary Table 4.** Goodness-of-fit statistics for excess male mortality during 2020. Legend: F P = F-statistic P-value, S.E. = standard error, SW P = Shapiro-WIlk P-value.

|  | F P | R² | S.E. R² | Δ* SW P | Δ SW P |
| --- | --- | --- | --- | --- | --- |
| Italy | 0.008 | 0.659 | 0.148 | 0.813 | 0.027 |
| Piemonte | 0.041 | 0.47 | 0.194 | 0.757 | 0.003 |
| Valle d’Aosta | 0.326 | 0.137 | 0.171 | 0.061 | 0.015 |
| Lombardia | 0.003 | 0.733 | 0.201 | 0.799 | <.001 |
| Bolzano | 0.002 | 0.778 | 0.105 | 0.065 | <.001 |
| Trento | 0.004 | 0.717 | 0.128 | 0.5 | <.001 |
| Veneto | 0.017 | 0.579 | 0.066 | 0.867 | 0.06 |
| Friuli Venezia Giulia | <.001 | 0.87 | 0.065 | 0.643 | 0.01 |
| Liguria | 0.008 | 0.661 | 0.147 | 0.657 | 0.015 |
| Emilia-Romagna | 0.01 | 0.632 | 0.156 | 0.752 | 0.007 |
| Toscana | 0.009 | 0.644 | 0.153 | 0.844 | 0.575 |
| Umbria | 0.008 | 0.654 | 0.149 | 0.113 | 0.189 |
| Marche | 0.081 | 0.372 | 0.205 | 0.023 | 0.027 |
| Lazio | 0.004 | 0.708 | 0.131 | 0.644 | 0.85 |
| Abruzzo | 0.012 | 0.62 | 0.16 | 0.239 | 0.004 |
| Molise | 0.01 | 0.637 | 0.155 | 0.529 | 0.274 |
| Campania | 0.023 | 0.544 | 0.18 | 0.236 | 0.182 |
| Puglia | 0.009 | 0.643 | 0.153 | 0.63 | 0.273 |
| Basilicata | 0.188 | 0.233 | 0.198 | 0.474 | 0.626 |
| Calabria | 0.015 | 0.594 | 0.167 | 0.338 | 0.395 |
| Sicilia | 0.038 | 0.482 | 0.192 | 0.23 | 0.58 |
| Sardegna | 0.003 | 0.749 | 0.116 | 0.744 | 0.056 |

**Supplementary Table 5.** Goodness-of-fit statistics for excess female mortality during 2020. Legend: F P = F-statistic P-value, S.E. = standard error, SW P = Shapiro-WIlk P-value.

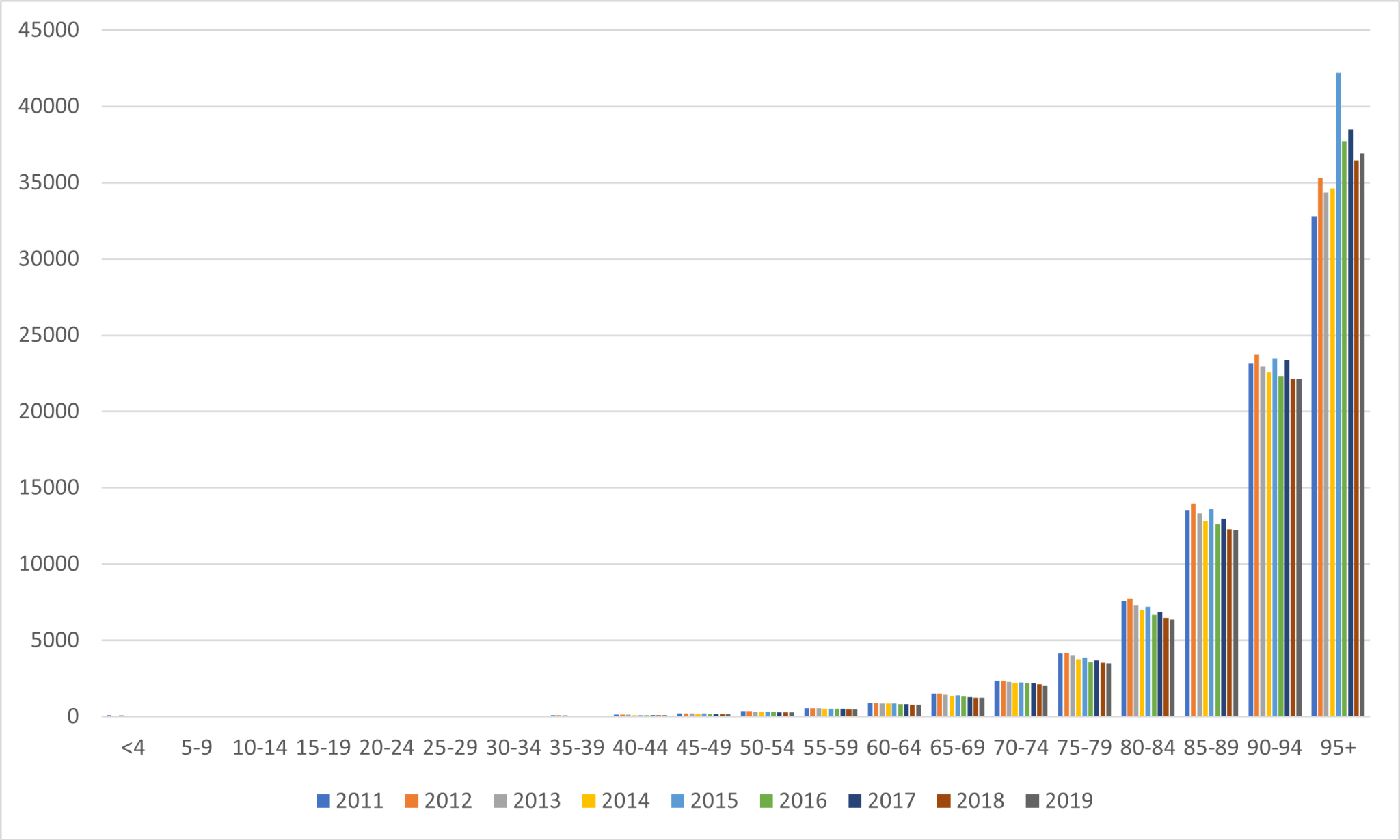

**Figure S1.** Male deaths per 100,000 inhabitants per age group from 2011 to 2019 (Italy).

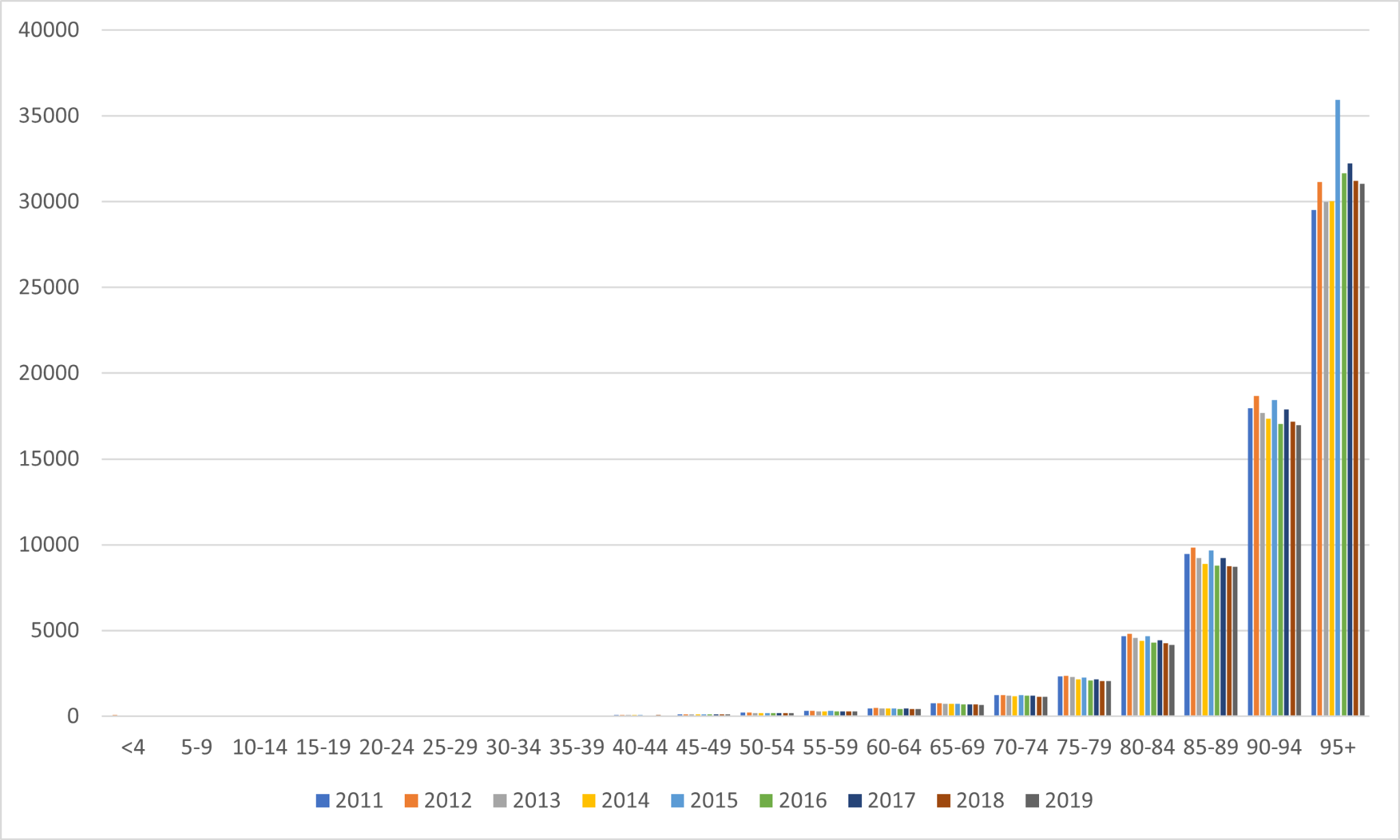

**Figure S2.** Female deaths per 100,000 inhabitants per age group from 2011 to 2019 (Italy).

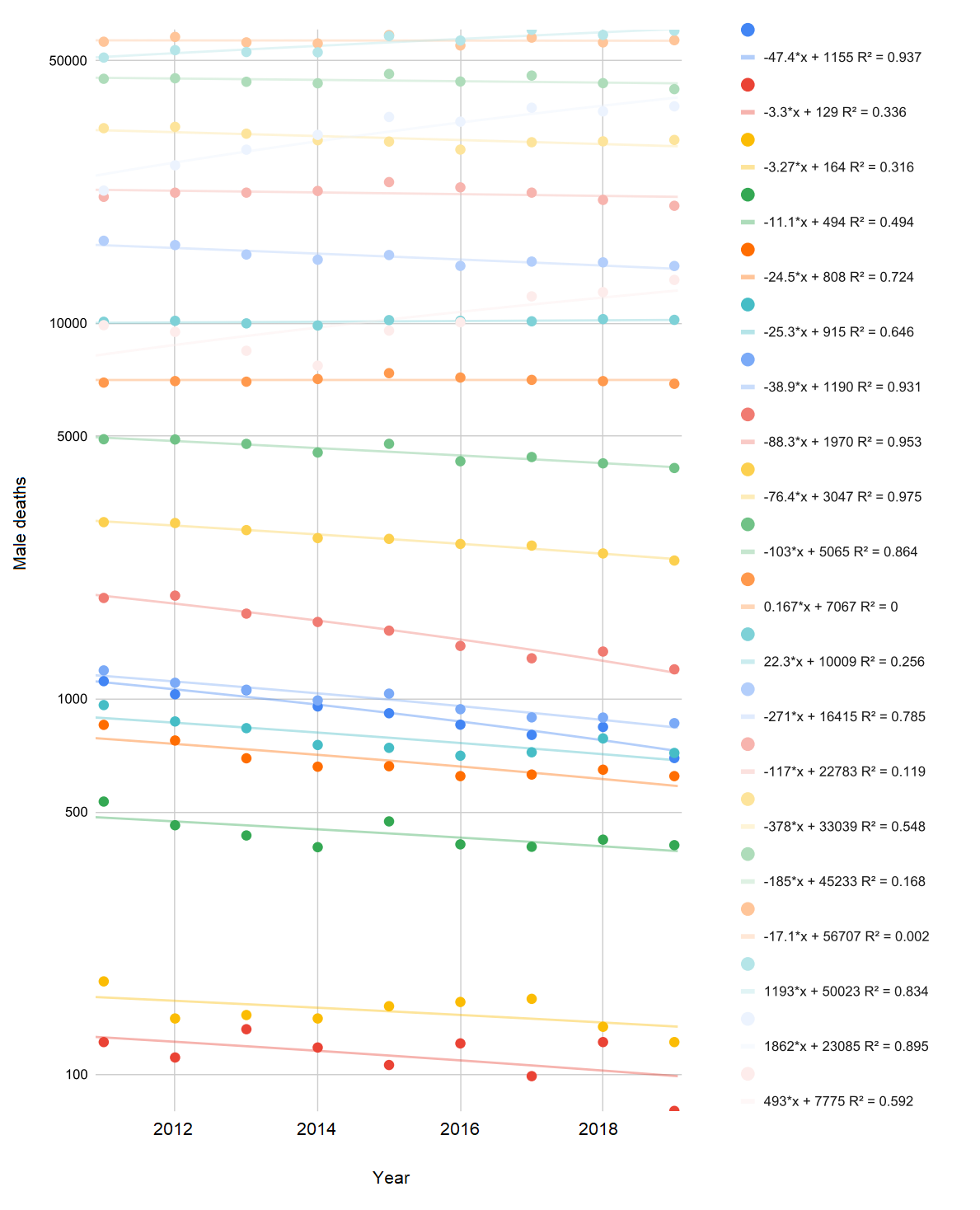

**Figure S3.** Male deaths per age group.

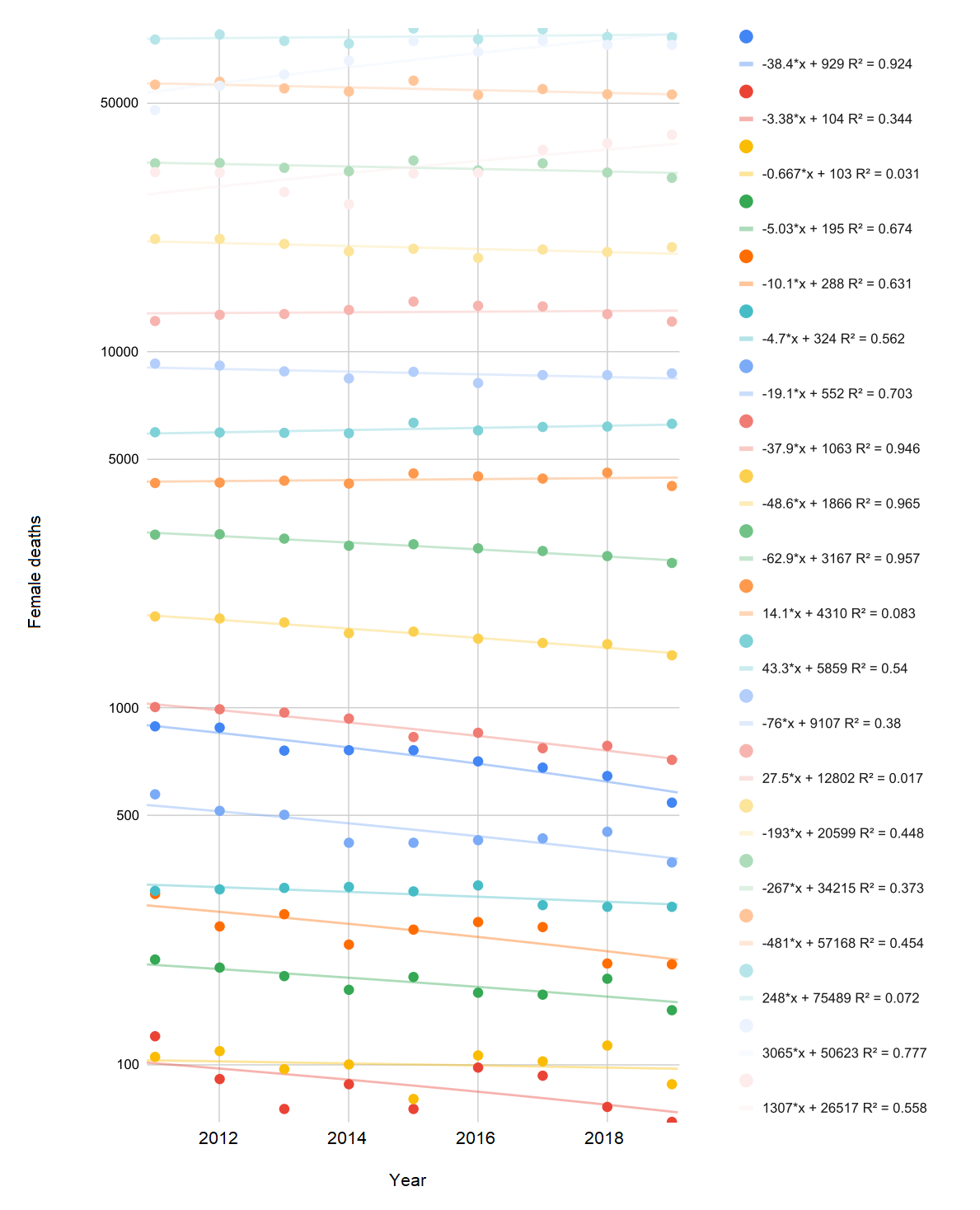

**Figure S4.** Female deaths per age group.
